# Contraceptive Method Skew in India 1992-2016

**DOI:** 10.1101/2020.07.14.20154013

**Authors:** Aalok Ranjan Chaurasia

**Author notes:** Address for correspondence: Aalok Ranjan Chaurasia, PhD, MLC Foundation and ‘Shyam’ Institute, 51, Lake City Farms (Ganeshpuri), Kalkheda Road, Neelbad, Bhopal, MP, India - 462044.

## Abstract

This paper analyses the contraceptive method skew in districts of India over more than two decades. The analysis reveals clear regional pattern in the method skew. In the northern and eastern regions, method skew is low and average but high or very high in the southern part of the country where factors such as poverty, education, social class, religion contributes little to deciding the method skew. In the central region, religious composition of the population has an impact on the method skew. In northern and eastern regions, poverty and education have an impact on the method skew. The analysis indicates that India has focussed on only one method - female sterilisation - to promote family planning, reduce fertility and curtail population growth. Increase in the prevalence of other family planning methods, especially, modern spacing methods, appears to be the need of the time. This requires improving the organisational efficiency and strengthening the administrative capacity of the official family planning services is the need of the time.

## Introduction

India is known for a contraceptive method-mix that is heavily skewed. The heavily skewed method-mix in India may be attributed to India’s preoccupation with reducing birth rate and curtailing population growth through birth limitation. During 1961-1971, India recorded an all time high population growth and, in response, the Government of India adopted a family planning based strategy of curtailing population growth. Specific demographic goal was set in terms of the desired birth rate which was translated into the number of new acceptors of different family planning methods to be recruited every year using demographic models. The rationale was to communicate some sense of urgency towards curtailing population growth in the country (Chaurasia and Singh, 2014). The focus of the strategy was on birth limitation while birth spacing was given on a residual attention. Official family planning efforts, therefore, promoted terminal methods of family planning - male and female sterilisation - with the aim of preventing 3rd and higher order births. This approach was, however, criticised on many grounds and an alternative reproductive and child health approach was suggested (Meesham and Heaver, 1996).

In 1996, the Government of India discontinued the target-based approach of family planning service delivery and introduced the target-free or the community needs assessment approach under the official National Family Welfare Programme. However, the emphasis on the promotion of terminal methods of contraception for birth limitation continued as curtailing population growth remained a priori development agenda. Fertility, in India, still remains above the replacement level (Government of India, 2017). The National Health Policy 2017 aims at reducing the total fertility rate to the replacement level and increasing the met need for family planning to more than 90 per cent at national and sub-national level by 2025 (Government of India, 2017a). A recent study has observed that the contraceptive method mix in India remains heavily skewed and there has been little change in the method mix in the country during 1992-2016 (Pradhan and Dwivedi, 2019). Other studies which have also demonstrated that the contraceptive method mix in India continues to be heavily skewed towards terminal methods of family planning, especially, female sterilisation, at the cost of modern spacing methods (Chaurasia and Singh, 2014; Ross, Keesbury and Hardee, 2015; Muttreja and Singh, 2018).

Studies on method skew in India have mostly been confined to country and state level. To the best of our knowledge, there has been no attempt to analyse the method skew across Indian districts. District level analysis of method skew is relevant as Indian districts are known for their strong social, cultural, economic and ethnic diversity. Variation in the method skew also reflects the variation in the quality of family planning services as it is well-known that a skewed method mix indicates limited contraceptive choice in terms of meeting the varied family planning needs of the people (Bertrand et al, 2000).

In this paper, we use the concept of the dominance of a particular contraceptive method over other methods to develop a method skew index (MSI) for analysing the method skew across Indian districts. The MSI is similar to the Herfindahl-Hirschman index in economics (Herfindahl, 1950, Hirschman, 1945; 1964), Simpson’s diversity index in ecology (Simpson, 1945), inverse participation ratio in Physics (Kramer and Mackinnon, 1993) and effective number of political parties index in politics (Laakso and Taagepera (1979). The next section defines the method skew index, MSI used in the present analysis. The third section of the paper describes data sources used for measuring and analysing contraceptive method skew across the districts of India. The analysis is based on the data available through nationally representative health and family welfare surveys that have been carried out in India since 1992. The fourth section presents the findings of the analyses at national and state/Union Territory levels. Inter-district variation in the method skew are analysed in section five. Section six of the paper applies the classification modelling approach to classify districts in terms of MSI after taking into consideration the selected defining characteristics of the district. The last section of the paper summarises the findings of the analysis and discusses their policy and programme implications in the context improving the quality of family planning services in the country through the promotion of a balanced method mix to meet the varied family planning needs of the people of the country.

## Method Skew Index

The contraceptive method skew has conventionally been measured in terms of the proportionate prevalence of different family planning methods available. Using this concept, the contraceptive method mix is termed as skewed if the proportionate prevalence of a single method is at least 50 per cent (Bertrand et al, 2015; Seiber, Bertrand and Sullivan, 2007; Sullivan et al, 2015). The limitation of this approach is that it categorises the contraceptive method mix in only two categories - skewed and not skewed. Ideally, method skew should be defined on a scale which ranges from equal proportionate prevalence for all contraceptive methods to proportionate prevalence of 1 for one method and 0 for all other methods available. Other approaches of measuring the contraceptive method skew are based upon comparing the observed method mix with a pre-specified standard. In this approach, method skew is measured in relative terms and not in absolute terms (Bertrand et al, 2000). Recently, method skew has been measured in terms of the average mean deviation (Ross, Keesbury and Hardee, 2015). The major limitation of this approach is that it also requires specification of the benchmark or the standard method. Since, family planning needs of the people are essentially dynamic in nature and preference for a particular contraceptive method is contingent upon a host of demand and supply side factors which are both endogenous and exogenous, specific of the benchmark is problematic. Moreover, if the standard or the benchmark is changed, the method skew measured in relative terms also changes.

In this paper, we measure the contraceptive method skew in terms of a method skew index (MSI) which is defined as the positive square root of the normalised weighted average of the proportionate prevalence of different contraceptive methods with weights being equal to the proportionate prevalence of the method itself. If *x*_i_ is the proportionate prevalence of method *I*, then method skew index (MSI) is defined as

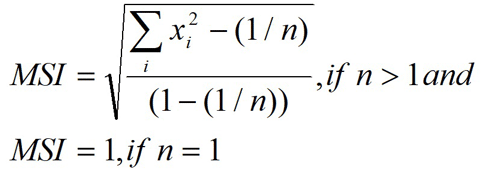

where *n* is the total number of contraceptive methods available. The MSI ranges from 0 through 1. When the entire contraceptive use is confined to only one contraceptive method, the proportionate prevalence is 1 for that method and 0 for all other methods available and, therefore, MSI=1. When the proportionate prevalence is the same for all the methods available, MSI= 0. MSI is independent of the number of contraceptive methods available. Based on the MSI, the contraceptive method skew may be classified as very low if MSI<0.200; low if 0.200≤MSI<0.400; average if 0.400≤MSI<0.600; high if 0.600≤MSI<0.800; and very high if MSI≥0.800.

There are a number of advantages in using MSI to measure and monitor the contraceptive method skew as compared to other approaches suggested. First, MSI measures the contraceptive method skew on a scale and not in specific categories as is the case with the 50 per cent rule. Second, it shows the absolute skewness in the method mix and, therefore, does not require specification of any standard or benchmark as is required when the method skew is measured in relative terms. Third, it is self-weighing in the sense that the higher the proportionate prevalence of a contraceptive method the higher the weight assigned to it and vice versa. As such, MSI is free from the bias that may be accrued by arbitrary selecting the weights. Fourth, it is the weighted sum of the proportionate prevalence and, therefore, is free from the problem of arithmetic compensability. Fifth, it is easy to calculate and can be monitored over time either on the basis of the survey data or even on the basis of the programme service statistics provided that the programme service statistics are of acceptable quality. Finally, the index does not require that the number of contraceptive methods available in the comparison space must be the same.

## Data

The analysis is based on the prevalence rates of different contraceptive methods for the country and for its constituent states/Union Territories and districts available from different rounds of National Family Health Survey (NFHS) and District Level Household and Facility Survey (DLHS) conducted by the Government of India from time to time during the period 1992-93 through 2015-16. The DLHS was a rapid household survey that was launched in 1998 as part of its Reproductive and Child Health Project which was later converted into Reproductive and Child Health Programme. Four rounds of DLHS were carried out - 1998-99 (Government of India, *no date*); 2002-04 (Government of India, 2006); 2007-08 (Government of India, 2010); and 2012-13. The first three rounds covered the entire country but the fourth and the last round was confined to selected states only. The DLHS was discontinued after 2012-13.

The NFHS, launched in 1992, is a large-scale, multi-round household survey to provide state and national level information on fertility, infant and child mortality, practice of family planning, reproductive and child health, nutrition, anaemia, utilisation and quality of health and family planning services, etc. Four rounds of NFHS have also been carried out since 1992-93 (Government of India, 1995; Government of India,2000; Government of India, 2007; Government of India, 2017b). The first three rounds provided estimates of method-specific contraceptive prevalence rate at the national and state/Union Territory level only. However, the fourth round provided estimates of method-specific contraceptive prevalence rates for each of the 640 districts in the country as they existed at the time of the survey. Estimates of the prevalence of six contraceptive methods - female sterilisation, male sterilisation, intra-uterine device, oral pill, condom and traditional methods - are available from these surveys for the period 1992-93 through 2015-16 for the country and states/Union Territories. The first five of these six methods constitute the basket of choice that is made available by the Government of India under the official National Family Welfare Programme. Method-specific prevalence rates for the districts, however, are available from 1998-99 onwards only. Recently, two more contraceptive methods - injectable and implant - have been added under the National Health Mission to expand the basket of choice of the contraceptive methods being offered to the potential beneficiaries of the Mission (Mutreja and Singh, 2018) but estimates of the prevalence of these two contraceptive methods are not available from any of the available sources either at the national level or at the local level. The data available from these surveys permit calculation of two variants of the method skew index (MSI). The first takes into consideration all the six contraceptive methods - modern and traditional. The second variant takes into consideration the modern contraceptive methods only. The method skew index taking into consideration all the six contraceptive methods is denoted by as MSI_a_ whereas the method skew index that takes into consideration only the five modern contraceptive methods is denoted by MSI_m_. Although, a substantial proportion of currently married women in India use traditional methods to regulate their fertility, yet these methods are neither supported nor promoted under the official family welfare programme.

## Method Skew in India and States/Union Territories

The method skew index taking into consideration all the six contraceptive methods (MSI_a_) increased marginally from 0.612 in 1992-93 to 0.619 in 2015-16 whereas the method skew index taking into consideration only modern contraceptive methods increased from 0.694 to 0.794 during this period. The trend in the method skew has, however, not been monotonous as MSI_a_ decreased during 1992-2004 but increased thereafter whereas MSI_m_ increased during 1992-99; decreased during 1998-2008 but increased, rather sharply, during 2015-2016 (Figure 1). It is, however, apparent from the data available through different nationally representative surveys that there has been little change in contraceptive method skew in the country during more than two decades between 1992-93 and 2015-16. It is also obvious that the major policy shift from a target-based approach to a community needs assessment approach in 1996 had only a marginal impact on the method skew and even this marginal impact appears to have waned in recent years.

**Figure 1:**
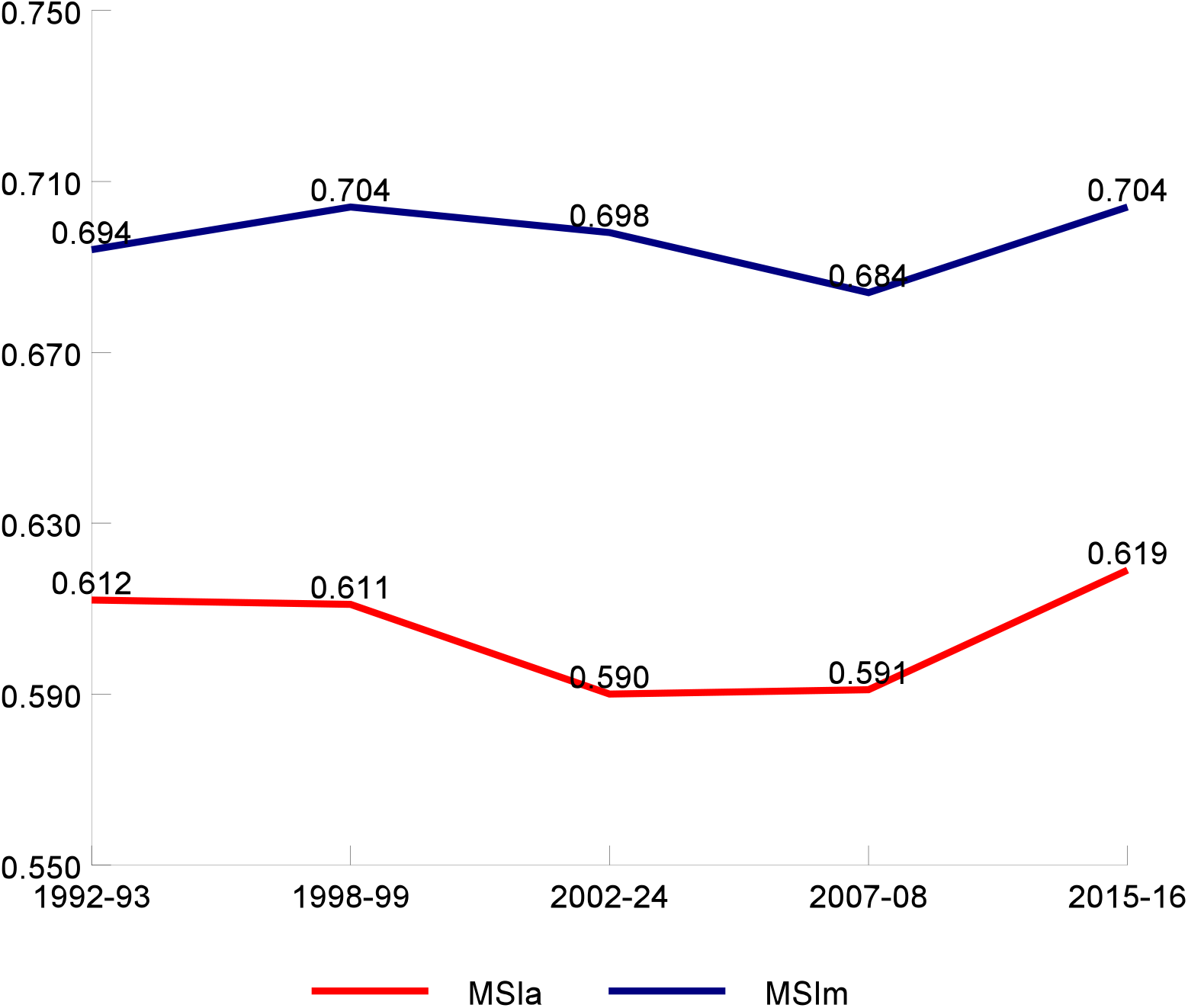
Trend in all methods skew index (*MSI*_*a*_) and modern methods skew index (*MSI*_*m*_) in India.

The method skew has also varied widely across the states/Union Territories of the country currently as well as in the past. There is no state/Union Territory where either MSI_a_ or MSI_m_ has been very low (less than 0.200) during the period under reference. On the other hand MSI_a_ is estimated to be very high (more than 0.800) in two states/Union Territories while MSI_m_ is very high in three states/Union Territories in 1992-93. These numbers increased to 7 and 8 respectively in 2015-16. The MSI_a_ was low in 3 states/Union Territories while MSI_m_ was low in 2 states/Union Territories in 1992-93 and these numbers also increase to 8 and 3 respectively in 1998-99. The a-divergence in the method skew is reflected in the increase in the inter-state/Union Territory coefficient of variation in MSI_a_ or MSI_m_. In 2015-16, the method skew was the highest in Andhra Pradesh in terms of both MSI_a_ and MSI_m_ but was the lowest in Nagaland in case of MSI_a_ and in Sikkim in case of MSI_m_. The north-south divide in the method skew is also very much evident from the table.

The trend in the method skew has also been different in different states/Union Territories. Out of the 25 states/Union Territories for which estimates of MSI_a_ and MSI_m_ are available from 1992-93 through 2015-16, MSI_a_ decreased in 12 states/Union Territories while MSI_m_ decrease in 14 states/Union Territories. The increase in both MSI_a_ and MSI_m_ has been the most rapid in Kerala. In the three southern states - Kerala, Tamil Nadu and Andhra Pradesh, both MSI_a_ and MSI_m_ increased quite rapidly. In Madhya Pradesh and Bihar also, both MSI_a_ and MSI_m_ increased very rapidly during the period under reference. By contrast, both MSI_a_ and MSI_m_ decreased very rapidly in Mizoram and Odisha. The strong state level effect on the trend in MSI_a_ and MSI_m_ is expected as the responsibility of the implementation of the National Family Welfare Programme actually rests with the constituent states/Union Territories. The state/Union Territory level preferences in the delivery of family planning services, therefore, dominate over national policy and programme priorities.

## Method Skew in Districts

The distribution of MSI_a_ and MSI_m_ across the districts of the country is given in Table 2. There is no district in the country during the period under reference where the method skew was very low (MSI_a_ or MSI_m_ < 0.200). Moreover, the proportion of districts having low method skew has decreased over time. On the other hand, the proportion of districts where the method skew is very high (MSI_a_ or MSI_m_, at least 0.800) has increased over time suggesting that the contraceptive use is getting more and more imbalanced in an increasing proportion of the districts. There are two districts in the country - Kurnool in Andhra Pradesh and Yadgir in Karnataka - where both MSI_a_ as well as MSI_m_ was 1 in 2015-16 which implies that the entire contraceptive use in these districts was confined to only one contraceptive method. The inter-district coefficient of variation in both MSI_a_ and MSI_m_, however, decreased between 1998-99 and 2015-16 which indicates convergence in method skew across districts. The regional distribution of MSI_a_ is depicted in figure 2 from which the north-south divide in the method skew is very much obvious. Districts with low or average MSI_a_ are primarily located in the northern and eastern parts of the country whereas MSI_a_ is very high in almost all districts in the southern part of the country. In Andhra Pradesh, Karnataka, Kerala and Tamil Nadu, the method skew is very high (MSI_m_ at least 0.800) in all but one districts. By contrast, there is only one district in the entire eastern part of the country where the method skew was very high in 2015-16. A similar situation prevails in the northern part of the country. In Bihar, Madhya Pradesh, Chhattisgarh and Jharkhand also, the method skew is found to be very high in a large proportion of districts.

**Table 1.**
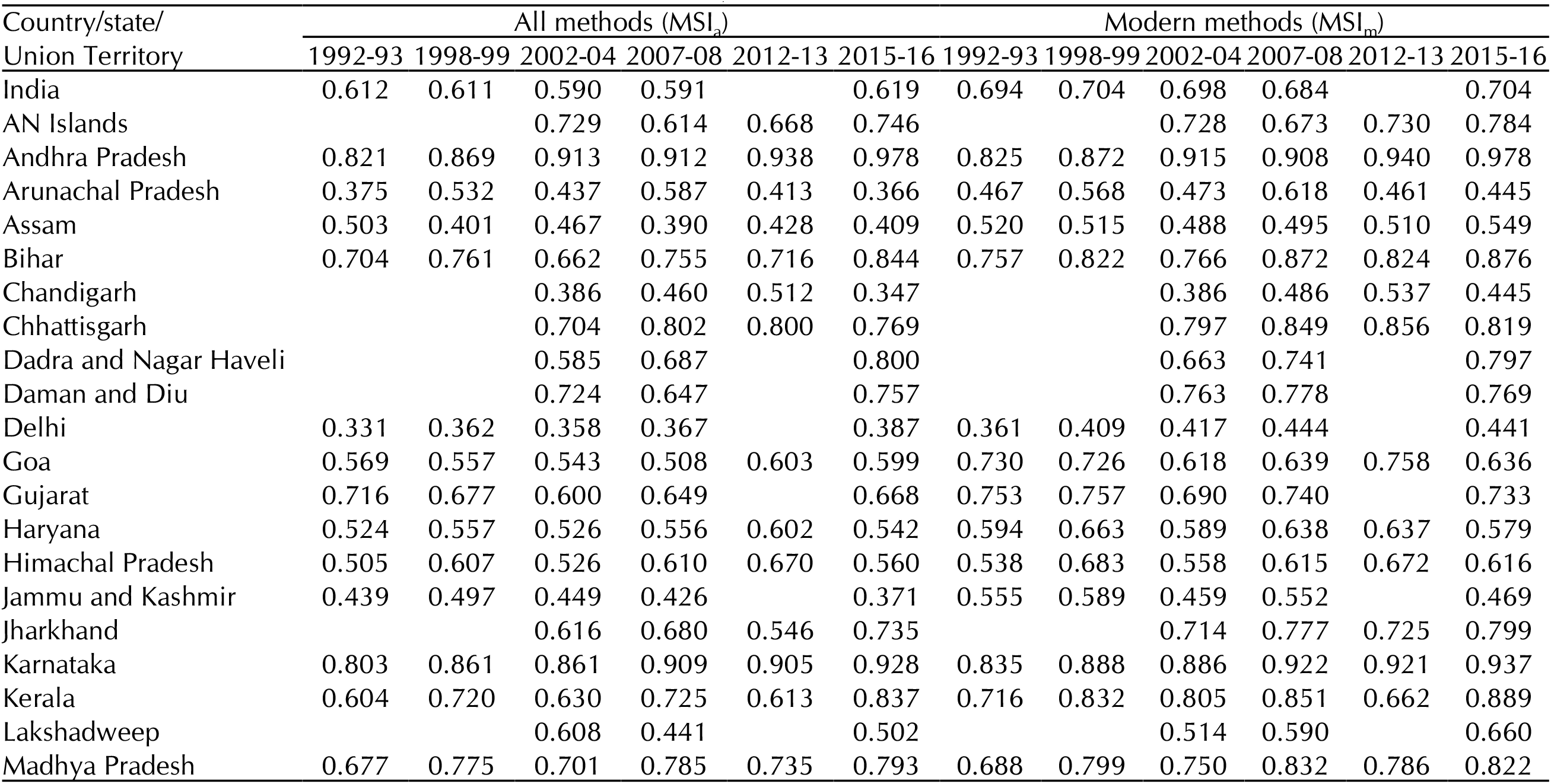
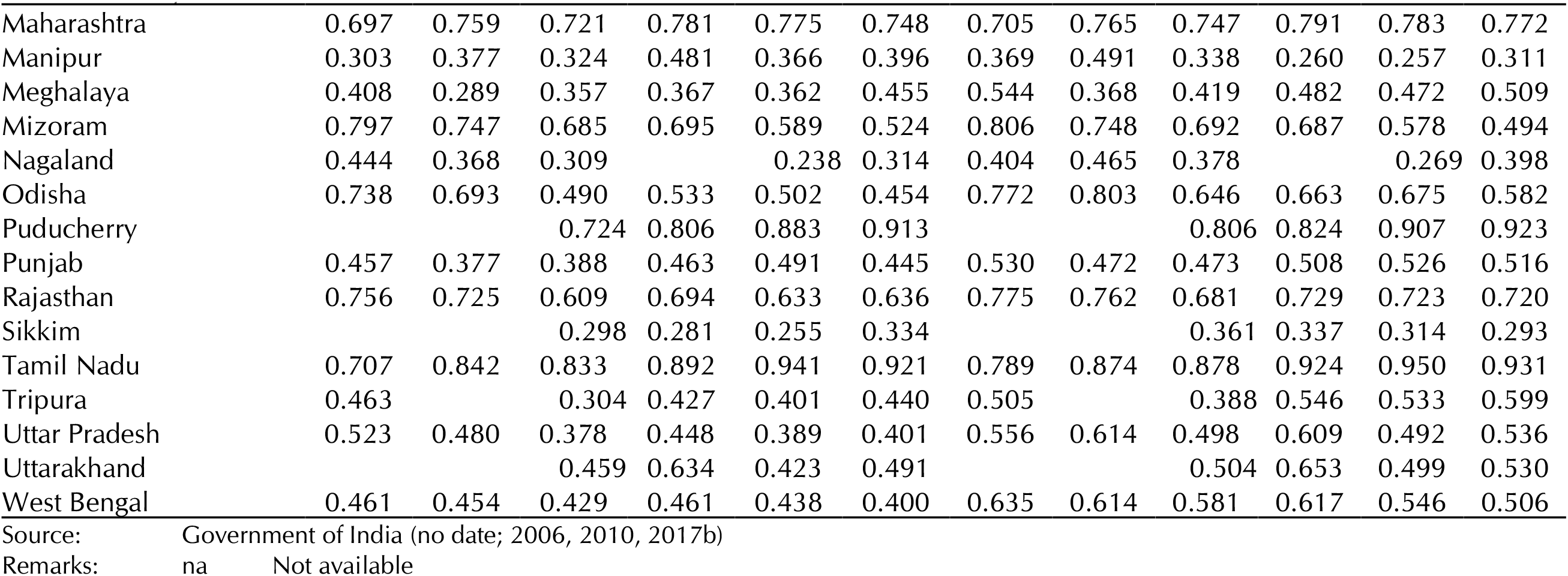
Method skew index (MSI) in India and states/Union Territories, 1992-2016

**Table 2.**
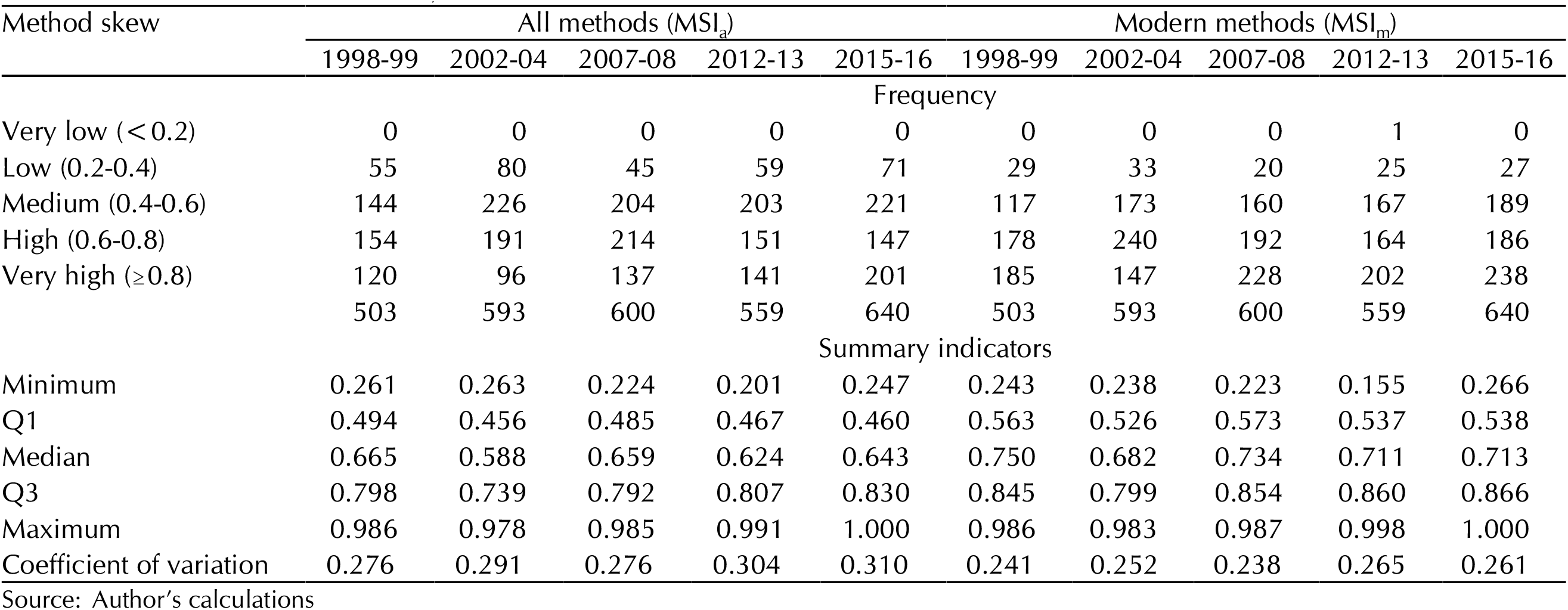
Inter-district variation in MSI in India, 1992-2016

**Figure 2:**
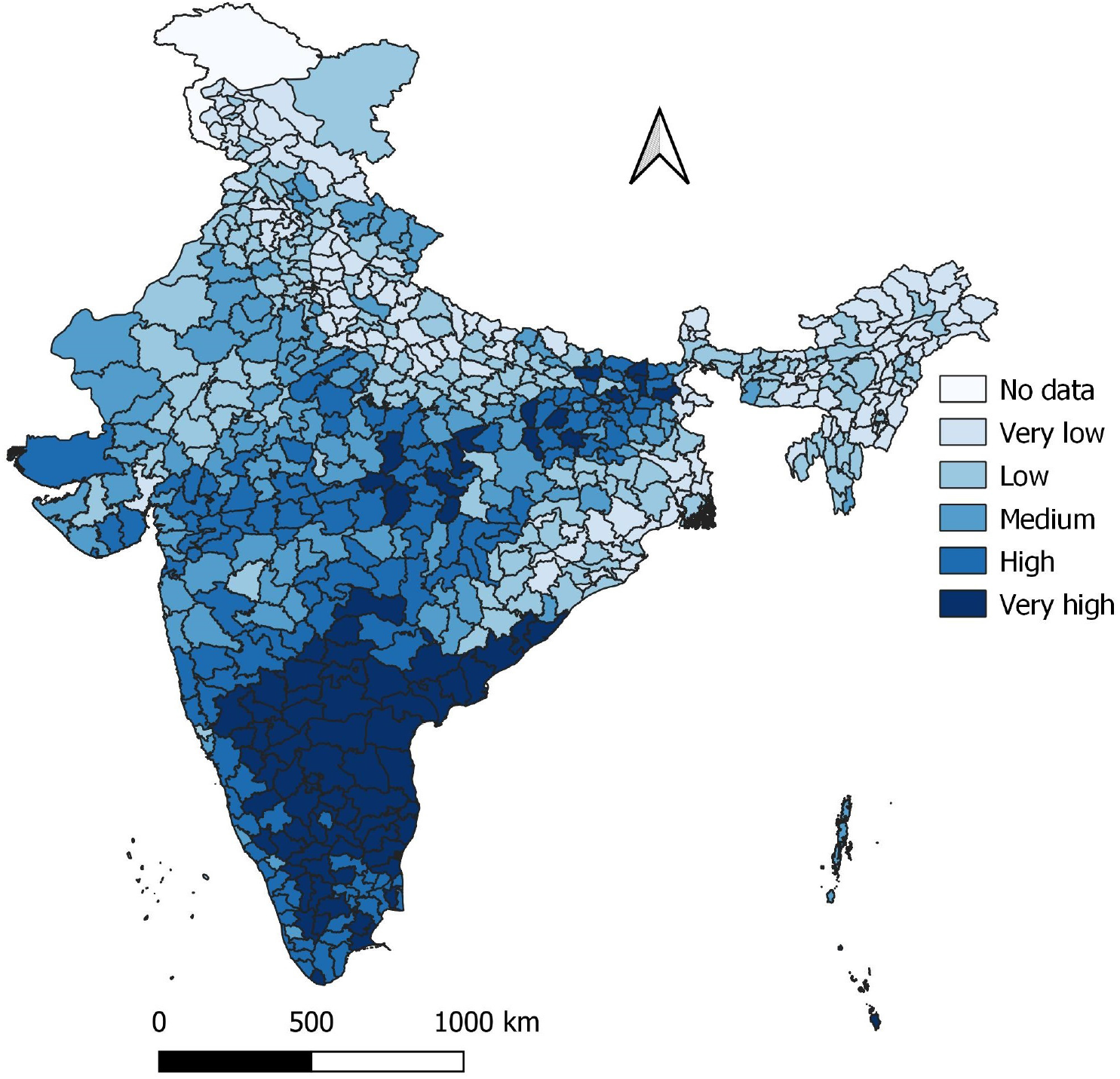
Inter-district variation in method skew index (*MSI*_*a*_) in India.

It has been hypothesised that a more equally distributed contraceptive method mix would be correlated with the contraceptive prevalence rate under the assumption that the expanded choice of contraceptive methods would lead to greater satisfaction and continuation in use and hence increase in the contraceptive prevalence rate (Bertrand et al, 2000).This means that the method skew index MSI_a_ or MSI_m_ should be inversely correlated with the contraceptive prevalence rate.

However, district level estimates of the contraceptive prevalence rate for the period 2015-16 negate this hypothesis. The Pearson product moment correlation coefficient between all methods contraceptive prevalence rate and MSI_a_ has been found to be positive but statistically insignificant (r=0.027). On the other hand, the Pearson product moment correlation coefficient between modern methods contraceptive prevalence rate and MSI_m_ has been found to be positive and statistically significantly (r=0.294). The positive correlation between method skew and contraceptive prevalence rate again confirms the dominance of a particular contraceptive method over others methods in majority of the districts of the country.

We have also counted the number of districts where the proportionate prevalence of any one of the six contraceptive methods is more than 50 per cent and the results are presented in table 3. The dominance of female sterilisation in the contraceptive methods mix in the districts can be judged from the observation that, during the period 2015-16, the proportionate prevalence of female sterilisation was more than 50 per cent in 438 or more than 68 per cent districts of the country, although the proportion of districts with proportionate prevalence of female sterilisation more than 50 per cent decreased from 1998-99 when the proportionate prevalence of female sterlisation was more than 50 per cent in more than three fourth districts. On the other hand, there are 14 districts where the proportionate prevalence of oral pill was more than 50 per cent. All these districts are located in the eastern part of the country. By contrast, the proportionate prevalence of IUD was more than 50 per cent in only one district (Leh in Jammu and Kashmir). Similarly, the proportionate prevalence of condom was more than 50 per cent in only one district (North East district in Delhi). Finally, there are nine districts where the proportionate prevalence of traditional methods of contraception has been found to be more than 50 per cent out of which six districts are located in Uttar Pradesh while the remaining three are located in Manipur, Assam and Jammu and Kashmir. At the same time, the number of districts where no method had a proportionate prevalence of more than 50 per cent has also increased over time from 110 (21.9 per cent) in 1998-99 to 177 (27.8 per cent) in 2015-16.

**Table 3.** Number of districts in which proportionate prevalence of a method is more than 0.500.

| Method | 1998-99 | 2002-04 | 2007-08 | 2012-13 | 2015-16 |
| --- | --- | --- | --- | --- | --- |
| Female sterilisation | 380 | 415 | 449 | 380 | 438 |
| Male sterilisation | 0 | 0 | 0 | 0 | 0 |
| IUD | 0 | 0 | 1 | 1 | 1 |
| Oral Pill | 1 | 1 | 2 | 3 | 14 |
| Condom | 1 | 3 | 0 | 1 | 1 |
| Traditional methods | 11 | 15 | 16 | 17 | 9 |
| No method | 110 | 159 | 132 | 157 | 177 |
| Number of districts | 503 | 593 | 600 | 559 | 640 |

Source: Author's calculations

## Classification of Districts

The method mix in a district is influenced by a host of factors including the orientation of the family planning services delivery system and factors related to the preferences and choices of the contraceptive users. Family planning services delivery in India has always been a prerogative of government efforts. As such, the orientation of the family planning services delivery has largely been shaped by the national policy towards family planning. However, the responsibility of implementing the national policy actually rests with the constituent states/Union Territories so that the state/Union Territory approach towards implementing the national policy has a major role in deciding the method mix in districts within the state/Union Territory concerned. In other words, the state/Union Territory factor may be argued to be an important factor in deciding the method skew at the districts level.

At the same time, it is well known that factors such as the extent of poverty, degree of urbanisation, level of education and religious and social class composition of the population, etc. also influence the individual method choice. These factors are exogenous to the family planning services delivery system. It is well known that these exogenous factors vary widely across the districts. It may, therefore be argued that a part of the inter-district variation in the contraceptive method mix may be attributed to the inter-district variation in these and other exogenous factors. An understanding of how these exogenous factors influence the contraceptive method mix at the district level is therefore important to understand the inter-district variation in the method skew.

We have followed the classification modelling approach (Han, Kamber, Pei, 2012; Tan, Steinbach, Kumar, 2006) to classify districts by the extent of the method skew in the context of the state factor, the extent of poverty in the district, the religious composition of the district population, the level of education and the degree of urbanisation in the district. The classification and regression tree (CRT) technique (Brieman, et al. 1982) was applied for the purpose. CRT is a nonparametric recursive partitioning method that divides districts into mutually exclusive groups in such a way that the within-group homogeneity with respect to the dependent variable is the maximum. A group in which all districts in the group have the same value of the dependent or classification variable is termed as “pure.” If the dependent variable is a categorical one, then the method provides the distribution of the dependent variable across all districts within the group. If the dependent variable is a scale or one, the method provides estimates of arithmetic mean and standard deviation of the distribution of the dependent variable across the districts within the group (Chaurasia, 2018).

For the purpose of classification, we have categorised the districts in four categories on the basis of the index MSI_m_ - districts where method skew was low (0.2≤MSI_m_<0.4) ; districts where method skew was average (0.4≤MSI_m_<0.6); districts where method skew was high (0.6≤MSI_m_<0.8); and districts where method skew was very high (MSI_m_≥0.8). On the other hand, the extent of poverty in the district was measured in terms of the proportion of households which did not have any of the seven household assets - radio/transistor, television, computer with or without internet, phone - landline or mobile or both, bicycle, any two-wheeler, and any four-wheeler. The level of education was measured in terms of the effective literacy rate which is defined as the proportion of the population aged seven years and above who can read and write with understanding; the degree of urbanisation was measured in terms of the proportion of the population living in the urban areas as defined at the 2011 population census; the religious composition of the population was measured in terms of the Muslim population in the district as proportion of the total population; and the social class composition of the population was measured in terms of the proportion of Scheduled Tribes and the proportion of Scheduled Castes in the population of the district. District level estimates of all the six independent variables for use in the classification modelling exercise were estimated from the data available through the 2011 population census.

Results of the classification modelling exercise are presented in table 4 and the resulting classification tree is depicted in figure 3. The exercise suggests that the 640 districts of the country can be classified into nine mutually exclusive groups or clusters having distinct characteristics in terms of the state/Union Territory in which the district is located, extent of poverty; the degree of urbanisation; proportion of Scheduled Castes population; proportion of Scheduled Tribes population and proportion of Muslim population in the district. The distribution of districts in terms of the method skew index MSI_m_ is different in different clusters. At the one end, there are 59 districts in the north and eastern states of the country where there is presence of the Scheduled Tribes population while the Scheduled Castes population was less than 1 per cent at the 2011 population census. In more than 93 per cent of these districts, the method skew was either low or average. In addition, there are 40 districts, also in the north and eastern parts of the country where the proportion of asset less households is less than 8.75 per cent at the 2011 population census. In more than 92 per cent of these districts, the method skew is average. At the other end, there are 109 districts, all located in the southern part of the country. In almost 95 per cent of these districts, the method skew is very high irrespective of the five independent variables included in the classification modelling exercise.

**Table 4.**
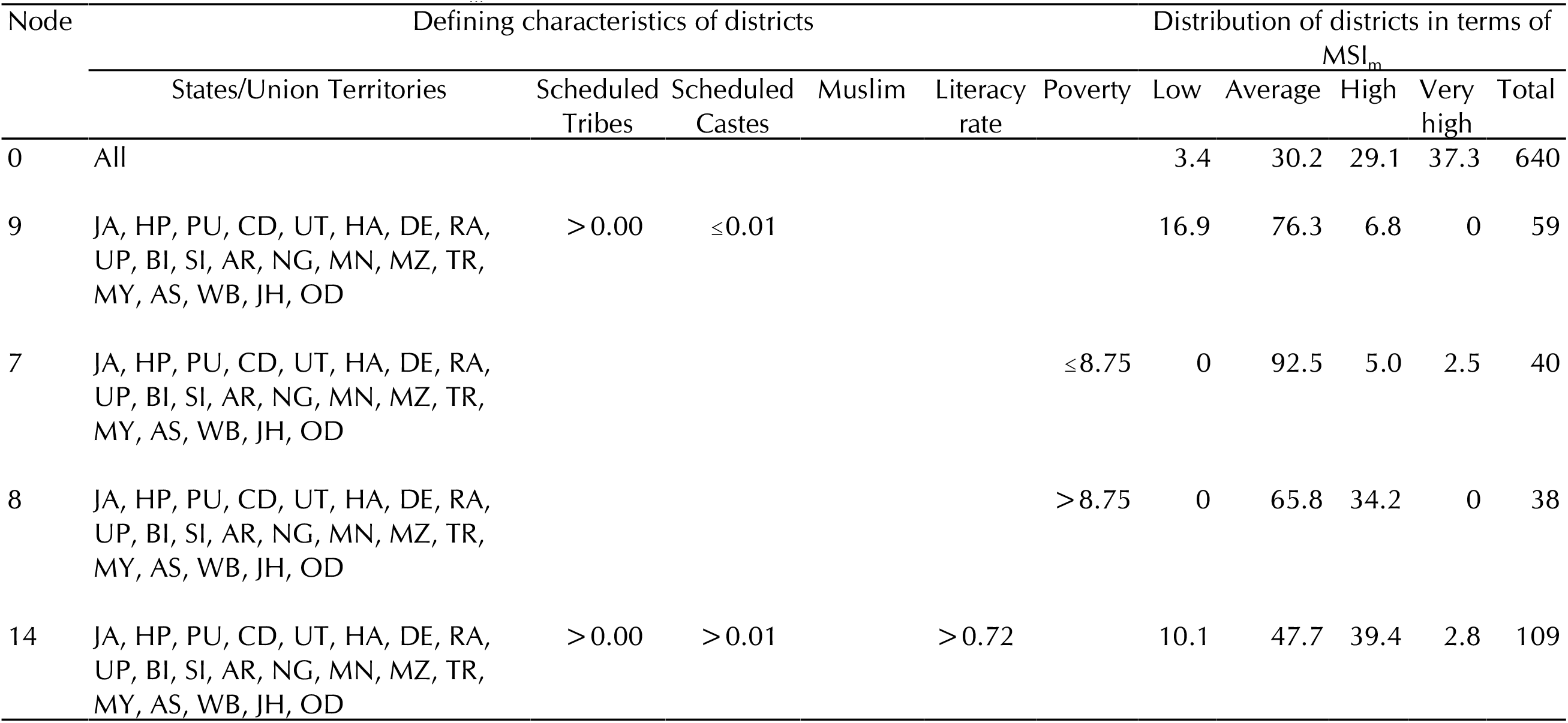
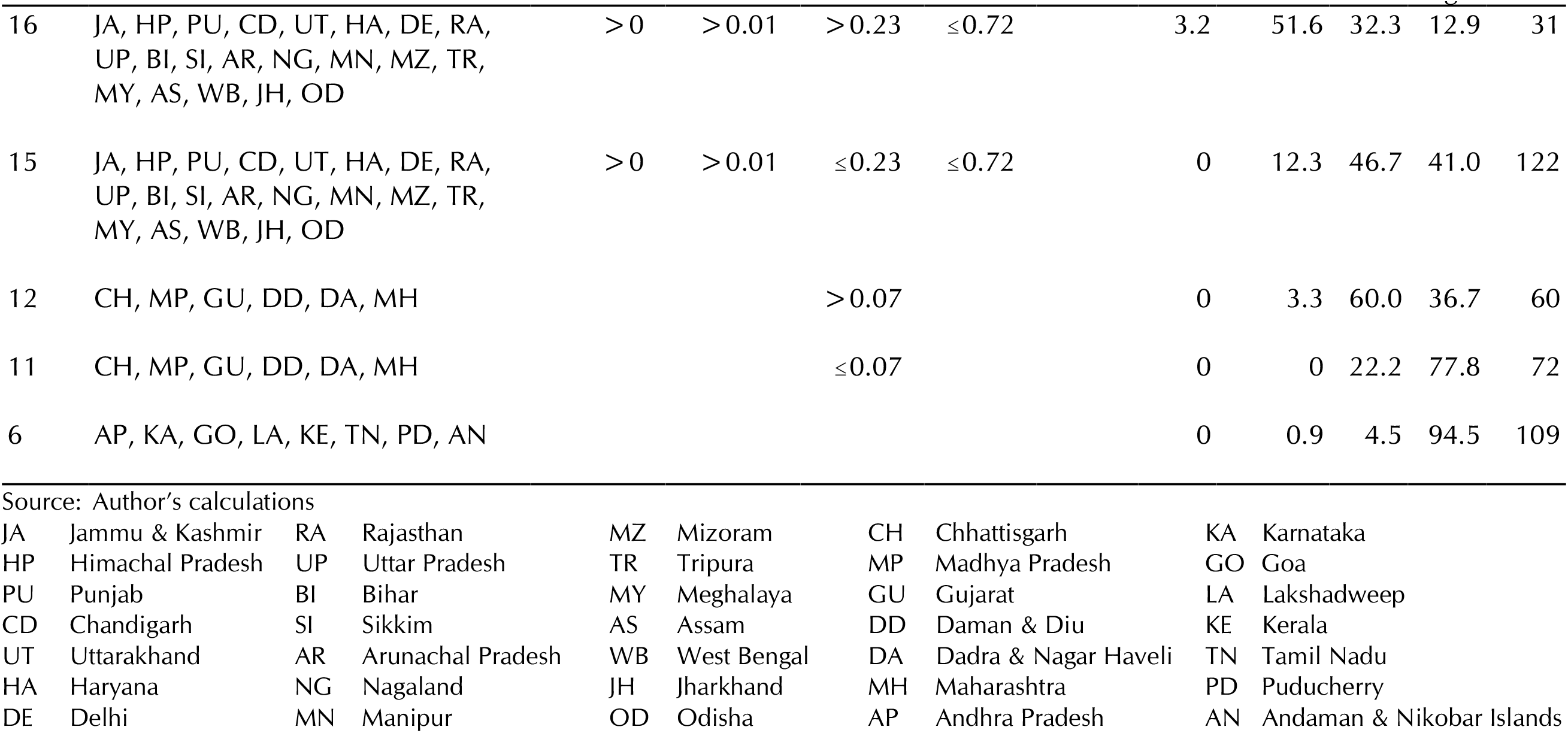
Classification of districts in terms of MSI_m_

| Node | Defining characteristics of districts | | | | | | Distribution of districts in terms of $MSI_m$ | | | | |
| --- | --- | --- | --- | --- | --- | --- | --- | --- | --- | --- | --- |
|  | States/Union Territories | Scheduled Tribes | Scheduled Castes | Muslim | Literacy rate | Poverty | Low | Average | High | Very high | Total |
| 0 | All |  |  |  |  |  | 3.4 | 30.2 | 29.1 | 37.3 | 640 |
| 9 | JA, HP, PU, CD, UT, HA, DE, RA, UP, BI, SI, AR, NG, MN, MZ, TR, MY, AS, WB, JH, OD | >0.00 | ≤0.01 |  |  |  | 16.9 | 76.3 | 6.8 | 0 | 59 |
| 7 | JA, HP, PU, CD, UT, HA, DE, RA, UP, BI, SI, AR, NG, MN, MZ, TR, MY, AS, WB, JH, OD |  |  |  |  | ≤8.75 | 0 | 92.5 | 5.0 | 2.5 | 40 |
| 8 | JA, HP, PU, CD, UT, HA, DE, RA, UP, BI, SI, AR, NG, MN, MZ, TR, MY, AS, WB, JH, OD |  |  |  |  | >8.75 | 0 | 65.8 | 34.2 | 0 | 38 |
| 14 | JA, HP, PU, CD, UT, HA, DE, RA, UP, BI, SI, AR, NG, MN, MZ, TR, MY, AS, WB, JH, OD | >0.00 | >0.01 |  | >0.72 |  | 10.1 | 47.7 | 39.4 | 2.8 | 109 |

| Node | Defining characteristics of districts | | | | | Distribution of districts in terms of $MSI_m$ | | | | | |
| --- | --- | --- | --- | --- | --- | --- | --- | --- | --- | --- | --- |
|  | States/Union Territories | Scheduled Tribes | Scheduled Castes | Muslim | Literacy rate | Poverty | Low | Average | High | Very high | Total |
| 16 | JA, HP, PU, CD, UT, HA, DE, RA, UP, BI, SI, AR, NG, MN, MZ, TR, MY, AS, WB, JH, OD | >0 | >0.01 | >0.23 | $\leq 0.72$ | | 3.2 | 51.6 | 32.3 | 12.9 | 31 |
| 15 | JA, HP, PU, CD, UT, HA, DE, RA, UP, BI, SI, AR, NG, MN, MZ, TR, MY, AS, WB, JH, OD | >0 | >0.01 | $\leq 0.23$ | $\leq 0.72$ | | 0 | 12.3 | 46.7 | 41.0 | 122 |
| 12 | CH, MP, GU, DD, DA, MH |  |  | >0.07 |  |  | 0 | 3.3 | 60.0 | 36.7 | 60 |
| 11 | CH, MP, GU, DD, DA, MH | | | $\leq 0.07$ | | | 0 | 0 | 22.2 | 77.8 | 72 |
| 6 | AP, KA, GO, LA, KE, TN, PD, AN |  |  |  |  |  | 0 | 0.9 | 4.5 | 94.5 | 109 |

Source: Author's calculations

|  |  |  |  |  |  |  |  |  |  |
| --- | --- | --- | --- | --- | --- | --- | --- | --- | --- |
| JA | Jammu & Kashmir | RA | Rajasthan | MZ | Mizoram | CH | Chhattisgarh | KA | Karnataka |
| HP | Himachal Pradesh | UP | Uttar Pradesh | TR | Tripura | MP | Madhya Pradesh | GO | Goa |
| PU | Punjab | BI | Bihar | MY | Meghalaya | GU | Gujarat | LA | Lakshadweep |
| CD | Chandigarh | SI | Sikkim | AS | Assam | DD | Daman & Diu | KE | Kerala |
| UT | Uttarakhand | AR | Arunachal Pradesh | WB | West Bengal | DA | Dadra & Nagar Haveli | TN | Tamil Nadu |
| HA | Haryana | NG | Nagaland | JH | Jharkhand | MH | Maharashtra | PD | Puducherry |
| DE | Delhi | MN | Manipur | OD | Odisha | AP | Andhra Pradesh | AN | Andaman & Nikobar Islands |

**Figure 3:**
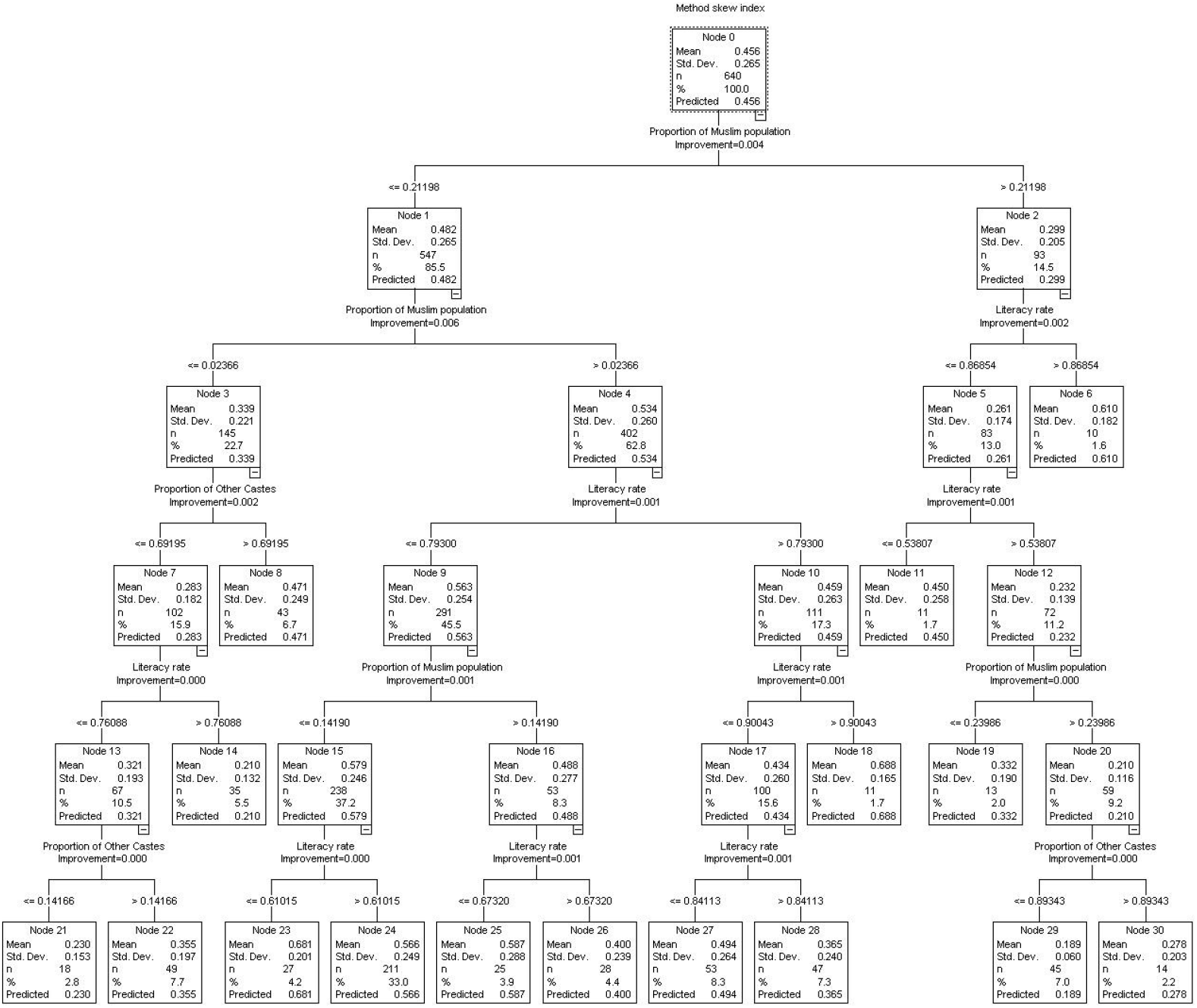
Classification of districts according to *MSI*_*a*_ and defining characteristics of the district.

**Figure 4:**
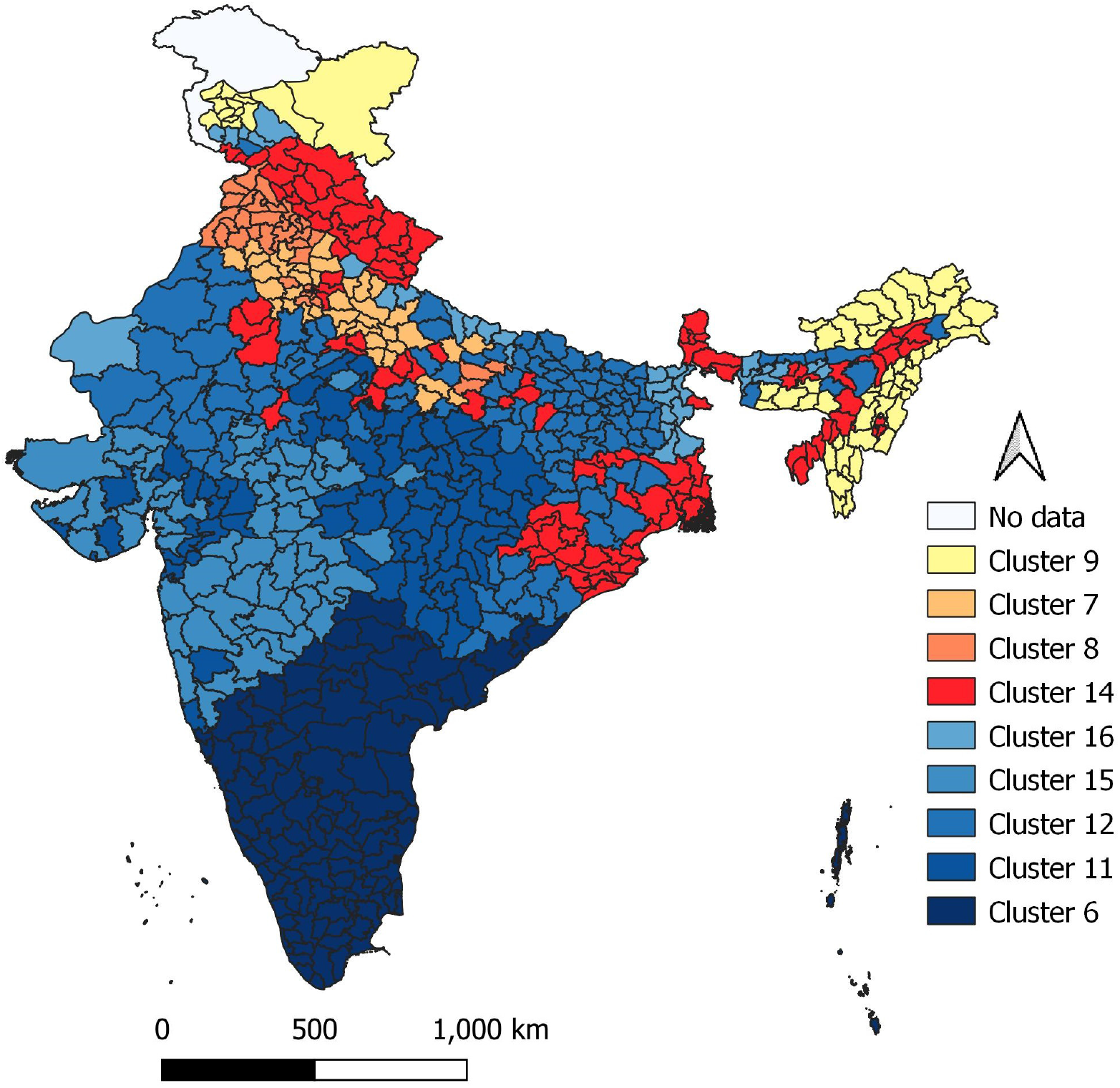
Cluster pattern across the districts of the country.

The classification modelling exercise suggests that India can be divided into three regions as regards method skew. The first region comprises of northern and eastern part of the country comprising of 21 states/Union Territories and 399 districts. All districts with low method skew and all but three districts having average method skew are located in this region. The second region comprises of six states/Union Territories located in the central part of the country and comprising of 132 districts. The method skew is very high in almost 60 per cent districts of this region. Finally, the third region comprises eight states/Union Territories, all located in the southern part of the country comprising of 109 districts of the country. The method skew is very high in almost 95 per cent districts of this region. The classification modelling exercise also suggests that in more than 38 per cent districts in the northern and eastern parts of the country where literacy rate is less than 72 per cent and where both Scheduled Castes and Scheduled Tribes are present, the proportion of Muslim population in the district has a strong impact on the contraceptive method skew. Similarly, in about one-fifth districts of this region, poverty, measured in terms of the proportion of asset less households influences the method skew. On the other hand, in the central part of the country, the proportion of the Muslim population plays a dominant role in deciding the method skew. In districts where the proportion of Muslim population is less than around 7 per cent of the district population, the method skew is very high in more than three-fourth of the districts. However, in districts where the proportion of Muslim population is more than 7 per cent, the method skew is very high in less than 40 per cent of the districts of the region. By contrast, in the southern part of the country, neither the social class composition of the population, nor the extent of poverty or the degree of urbanisation or the effective literacy rate or the religious composition of the population appears to have any significant role in deciding the contraceptive method skew because the contraceptive use in this part of the country is virtually confined to only one method of contraception - female sterilisation.

## Discussions and Conclusions

The present analysis is probably and so obviously the first to examine method skew at the district level in India. The analysis is important because family planning services in India are essentially planned and delivered at the district level, although, the policy towards family planning is laid down at the national level and customised at state/Union Territory level. The analysis is based on a new index which is based on the concept of the dominance of one method over others. The index used in the present analysis has a number of advantages over other approaches used to measure the method skew - the 50 per cent rule and the average deviation approach. The 50 per cent rule categorises the contraceptive method mix in only two categories - skewed or not skewed - whereas the average deviation approach requires specification of a standard. The index does not changes with the change in the number of contraceptive methods available. It can serve as a useful tool to measure and monitor the orientation of the family planning services delivery system in the context of the felt family planning needs of the community.

The analysis presented in this paper suggests little change in the method skew in India over the last two decades and more. Moreover, the method skew in the districts of the country is directly related to the contraceptive prevalence rate, particularly, the prevalence of female sterilisation - the higher the prevalence of female sterilisation the higher the contraceptive prevalence rate and the higher the method skew. Although, at the policy level, the government professes availability of a basket of family planning methods to meet the varied family planning needs of couples, yet the high to very high method skew in most of the districts of the country suggests that, in reality, the choice of family planning methods is severely restricted.

The analysis also suggests that there is clear regional pattern across the districts of the country as far as the method skew is concerned. In the northern and eastern parts of the country, the method skew is low and average in most of the districts whereas in all districts of the southern part of the country, the method skew is high or very high. In districts located in the southern part of the country, factors such as the extent of poverty, level of education, and social class and religious composition of the district population contributes little to deciding the method skew because of the near universality of high to very high method skew. In the central part of the country, however, the religious composition of the population do have an impact on the method skew. On the other hand, in the northern and eastern parts of the country, the extent of poverty and the level of education appear to have an impact on the method skew.

India was able to increase the contraceptive prevalence rate from around 36 per cent in 1992-93 to almost 55 per cent in 2007-08 by focussing on only one method of family planning - female sterilisation - in the quest towards limiting the number of births per couple thereby reducing the fertility and curtailing population growth. However, the contraceptive prevalence rate has decreased to around 53 per cent in 2015-16. This shows that continued emphasis on only one method of contraception, especially, female sterilisation, may not be the right approach to meeting the future family planning needs of the Indian population. Increasing the prevalence of other family planning methods, especially, modern spacing methods, appears to be the need of the time. However, organisational efficiency and administrative capacity of the official family planning services delivery system which is the mainstay of family planning efforts in the country, remains weak in this context.

## Data Availability

All data are available openly.

